# Increase of non-vaccine human papillomavirus types in a group of HPV-vaccinated Mexican women. Evidence of Pathogenic Strain Replacement

**DOI:** 10.1101/2020.05.26.20113829

**Authors:** Augusto Cabrera-Becerril, Ramiro Alonso, Cruz Vargas-De-León, Pedro Miramontes, Pablo Romero, Keiko Taniguchi, Daniel Marrero, Marco Capistrán, Sheila Jiménez, Sonia Dávila, Mauricio Salcedo, Raúl Peralta

**Affiliations:** Departamento de Matemáticas, Facultad de Ciencias, Universidad Nacional Autónoma de México; Centro de Investigación en Dinámica Celular, Universidad Autónoma del Estado de Morelos; Escuela Superior de Medicina, Instituto Politécnico Nacional; Unidad de Investigatión Médica en Enfermedades Oncológicas, Instituto Mexicano del Seguro Social; Centro Médico Universitario, Universidad Autónoma del Estado de Morelos

**Keywords:** vaccine HPV, non-vaccine HPV, Vaccine-induced Pathogen Strain Replacement, SIR model

## Abstract

High risk HPV infection is the etiological factor of Cervical Cancer (CC) and other types of cancer of epithelial origin. HPV 16 and 18 infections are associated with 70% of CC worldwide. At the present time, there is a vaccine that prevents this infections. In Mexico, the HPV vaccine was introduced in 2009. Even if the current vaccine is effective, some models indicate a possible scenario of Vaccine-induced Pathogen Strain Replacement (VPSR). In this report, we performed the molecular detection of HPV in a group of HPV-vaccinated Mexican women to explore a putative scenario of VPSR. We used biological samples from women who went for their routine Pap. The study included eighteen women older than 18 years of age and HPV-vaccinated. As the number of cases analyzed is relatively small, we supplemented the study with an agent-based direct computer simulation. The outcome of the numerical experiments and the analyzed cases complement each other and show that in three different scenarios, there is an increase in HPV cases approx 10 years after vaccination of the first cohort of women. The prevalence of non-vaccine HPV types increases when compared to prevalence of vaccine HPV types. This result could be interpreted as the phenomenon of Vaccine-induced Pathogenic Strain Replacement.

## 1 Introduction

Human Papillomavirus (HPV) infection is the causative biological agent of Cervical Cancer (CC) and other types of cancer of epithelial origin. This virus family includes more than 200 HPV types or genotypes, among which 40 genotypes infect mucous tissues such as the genital tract. Fourteen HPV types (HPV 16, 18, 31, 33, 35, 39, 45, 51, 52, 56, 58, 59, 66, and 68) are considered as highrisk (HR) oncogenic for causing CC development. This finding has driven the development of vaccines that help preventing HPV16 and HPV18 infections. Both genotypes are associated with 70% of CC worldwide [1]. In Mexico, the HPV Vaccination Program began in 2009 using the tetravalent vaccine, which protects against HPV types 6, 11, 16, and 18, which are the main causes of most genital warts and of CC. This vaccination scheme is aimed at girls aged 9-14 years before sexual initiation [2].

The current vaccine is effective. However, it is possible that it could cause an increase in the prevalence of infections with different genotypes through Vaccine-induced Pathogenic Strain Replacement (VPSR). Some studies have reported that the HPV vaccine could confer cross-immunity against related HPV genotypes but, on the other hand, according to an ecological notion, the removal of HPV dominant (HPV 16 and 18) as the result of vaccination may result in an increase in transmission of the HPV not dominant. Also, it is expected that with the application of the nonavalent vaccine the VPSR phenomenon does not occur. This last vaccine is still not applied by the Secretariat of Health in Mexico [3].

Several studies report the increase in non-vaccine infections in women who have been HPV-vaccinated; for example, in a follow-up study in Scotland, Cameron et al. reported a decline in vaccine HPV16 and 18, a significant decrease in non-vaccine HPV31, 33, and 45 (suggesting cross-protection), and a non-significant increase in non-vaccine HPV51 [4]. In another study conducted in England, it was found that the national HPV Immunization Program is successfully preventing HPV16/18 infection in sexually active young women. The detected prevalence of non-vaccine hr HPV types was slightly higher during the post-immunization period. These increases, combined with the decreases in HPV16/18, resulted in a similar prevalence of all hr HPV i.e., vaccine and non-vaccine types [5]. Similar results in the Australian HPV Vaccination Program, that is, a substantial fall in vaccine HPV types in HPV-vaccinated women and a lower prevalence of vaccine HPV types in unvaccinated women, suggested herd immunity and a possible indication of cross-protection against HPV types related to vaccine HPV types in HPV-vaccinated women [6]. Similar results found in Sweden, that is, a major reduction of HPV6, 16, and 18 prevalence and the minor changes observed for non-vaccine HPV types will require further investigation [7]. A recent sudy examined the cross-protection and type replacement after HPV vaccine and determined changes in proportions of non-vaccine HPV types prevalence and shows evidence of cross-protection with genetically related vaccine HPV types. The authors do not discard the possibility of type replacement for non-vaccine HPV types not genetically related.[8]

In order to define a possible mechanism of VPSR, in a previous work [9], our group examined the HPV dynamic after actual vaccination scheme employing a mathematical Susceptible, Infected, and Recovered (SIR) epidemiological model now we use it for normalize the experimental data obtained of a group of Mexican women HPV-vaccinated and from the results of the normalized data we setup three experimental scenarios for the agent-based model proposed in the next section.

## 2 Methods

### 2.1 Biological samples and DNA purification

This study was approved by the Local Research Committee of University Health Center of the Morelos State University (UAEM). All methods in this study were carried out in accordance with relevant guidelines and regulations. All participants in this study signed an informed consent and waived biological samples for this study. Data and biological samples were collected from women who attended for their routine Pap test at the University Health Center of the Morelos State University (UAEM) located in Cuernavaca, Mexico, during the 2016-2017 period. The study included any women older than 18 years of age, who were HPV-vaccinated. Cervical scrapes were obtained with cytobrush from women with normal cervix. For DNA extraction, the Wizard Genomic kit (Promega, Madison, WI, USA) was used according to the manufacturer’s instructions. DNA was purified, then quantified in a NanoDrop Spectrophotometer ND-1000 and resolved in 1% ethidium bromide-stained agarose gel.

### 2.2 HPV genotyping

The samples were genotyped by a microarray test according to manufacturer’s protocol (Genomica, Madrid, Spain). We utilized the HPV2 ® Clinical Array kit and employed only 10 *μ*g of purified DNA for each specimen. The kit utilizes biotinylated primers to define a sequence of 451 nucleotides within the polymorphic L1 region of the HPV genome. A pool of HPV primers is used to amplify HPV DNA from 35 mucosal genotypes, including hr HPV (16, 18, 31, 33, 35, 39, 45, 51, 52, 56, 58, 59, 68, 73, and 82) types, three potentially hr HPV (26, 53, and 66) types, 11 Low-Risk (lr) HPV (6, 11, 40, 42, 43, 44, 54, 61, 70, 72, and 81) types, and six of Unknown-Risk (ur) HPV (62, 71, 83, 84, 85, and 89) types. An additional primer pair that targets the human Cystic Fibrosis Transmembrane Conductance Regulator (CFTR) gene was employed as a DNA control for cell adequacy, extraction, and amplification. This primer pair also amplifies a fragment of CFTR located on a modified plasmid internal control, thereby avoiding false-negative results due to an inadequate PCR. If neither of the controls is positive, the specimen is considered inconclusive. The detection system is based on the precipitation of an insoluble product in zones of the microarray in which the hybridization of the amplified products with the specific probes takes place. During PCR, the amplified products are labeled with biotin. After amplification, these products hybridize with their respective specific probes, which are immobilized in known and particular areas of the microarray, after which the product is incubated with a Streptavidin-peroxidase conjugate. The conjugate binds via Streptavidin to the biotin present in the amplified products and the peroxidase activity gives rise to the appearance of an insoluble product in the presence of the substrate o-Dianisidine, which precipitates on the areas of the microarray in which hybridization occurs (Figure 1).

**Figure 1:**
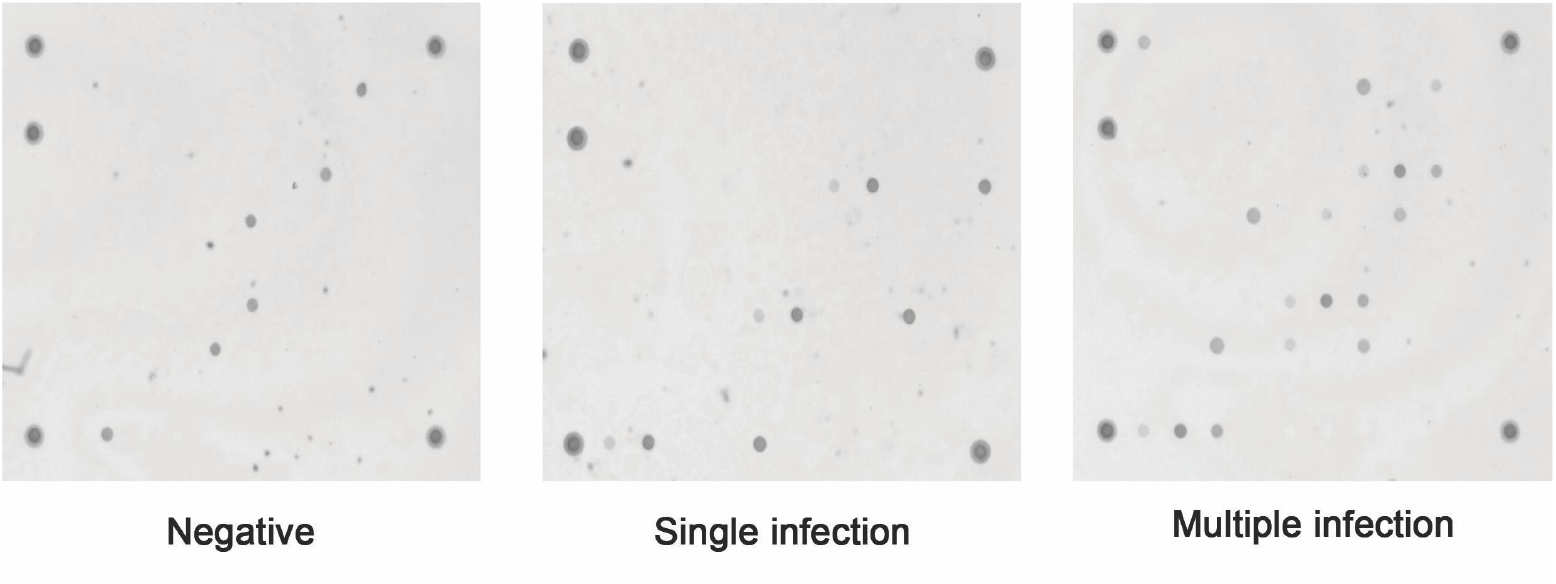
Visualization of the genotyping system of the CLART HPV2 system. Each point represents the hybridization of the genome of a type of HPV with its targeting the micro-array. Big spots over the edges of the square are control points. Interpretation of the results are provided by the system software

### 2.3 Statistical analysis

Clinical association analysis was carried out by means of the Fisher exact test. All *p* values represented two-tailed tests and were considered significant at < 0.05, and with the calculated *p* value using 100, 000 bootstrap samples. Statistical analysis was performed utilizing the SPSS ver 15 statistical software program, with the exception of the bootstrap that was performed in R statistical software.

### 2.4 Computational model

Mathematical and computational modeling in medicine has a long tradition of mutual benefit between mathematics and medicine. It is worth mentioning the classical cases of the action potential Hodgkin-Huxley model [10] and the more recent success of artificial neural network models [11]. An notorious case of mathematical modeling support arises when wet lab experiments are difficult or even impossible due to financial or ethical reasons. Traditionally, differential equations have been used as mathematical models in medicine. This implies, a relatively large number of parameters with the concomitant problem of finding their values. Although this approach has been useful, the advent of digital computers have opened the possibility of making direct modeling. Namely the process of mimicking the studied phenomenon directly with few, if any, assumptions and parameters. In our case, we admit that the number of women analyzed is small for a clinical study. We accordingly complement our study with a mathematical model designed to closely incorporate the main traits of the vaccination phenomenon.

Agent-based modeling (ABM) stands for a class of computational models that describes the dynamics of a many particles system using individual interactions. ABM has been used for the modeling of cancer systems in “HPV-VaC-min” is an agent based-model which search for possible emergent behavior on a sex-structured population. A certain fraction of this population have been infected with HPV. We have studied the infectious dynamics of HPV with vaccination scheme as a control mechanism. Here we use a reduced implementation of “HPV-VaC” model described in [26]. on the NetLogo [28] framework. We ran several numerical experiments described in Table 1. The code for the NetLogo implementation for this model can be consulted in [27]. Afterwards we analyze the data collected in numerical experiments using python language.

**Table 1:** Variables of the model.

| Variable | Type | Description |
| --- | --- | --- |
| Probability of infection | Global | Average rate of transmission |
| Symptoms show | Global | Average time for an infected to show any symptoms |
| Initial population | Global |  |
| Recruitment rate | Global |  |
| hline Rate of vaccination | Global |  |
| Couple life time | Global | Average time of couple relationship duration |
| Subpopulation | Agent | Sane , Infected, Sick , Cin-1,2,3 , Vaccinated, diagnosed |
| Virus | Agent |  |
| Infection time | Agent |  |
| Social Status | Agent | single or coupled |
| Coupling tendency | Agent |  |
| Couple duration | Agent | Average time for the couple relationship. |
| Age | Agent |  |
| Promiscuity index | Agent |  |
| Coupling effectiveness | Agent |  |
| Disease stage | Agent | Cin-1,2,3 CCU |
| Vaccine Duration | Agent | Random number between 5 and 7 |

Based on the literature and our previous estimations in [9] we assume as hypothesis for the model that

1. The probability of infection is distributed normally over the population.[9]
2. The chance of recovery is distributed as a weighted normal over the population, the weights decrease as the disease progresses from CIN1 to CIN2 and CIN3.[9]
3. Primary infection is asymptomatic, there is a latency period that varies from 2 weeks to 10 years.[12]
4. Active sexual life is taken from 13 to 60 years of age.[9]
5. Women take the pap test once over three years.[9]
6. Vaccinated Women increases sexual activity.[13]
7. The vaccine loses effectiveness after 5 to 10 years.[14]
8. The vaccine offers cross protection to hpv types genetically related in the same species, so we consider the pentavalent vaccine essentially a bivalent vaccine and the 9-valent vaccine a 3-valent vaccine[8]
9. Rate of infection depends strongly on the sexual collective behaviour of population.[13]

In this model (HPV-VaC) we have two kind of variable: global and agent. Global variables are control parameters and agent variables are state variables for each individual agent. In Table 1 we summarize description of global and agent variables.

The interaction space between agents it’s a 101 × 87 square grid with boundary conditions of a flat torus. The implementation on the NetLogo framework has two basic methods: **Setup** and **Go**. **Setup** method have two functions: **Setup_people** and **Setup_globals**. **Setup_globals** initialize global variables. **Setup_people** initialize agent state-variables. This two functions set the space of parameters and give certain agent heterogeneity. We have depicted the **Setup** method on Figure 2

**Figure 2:**
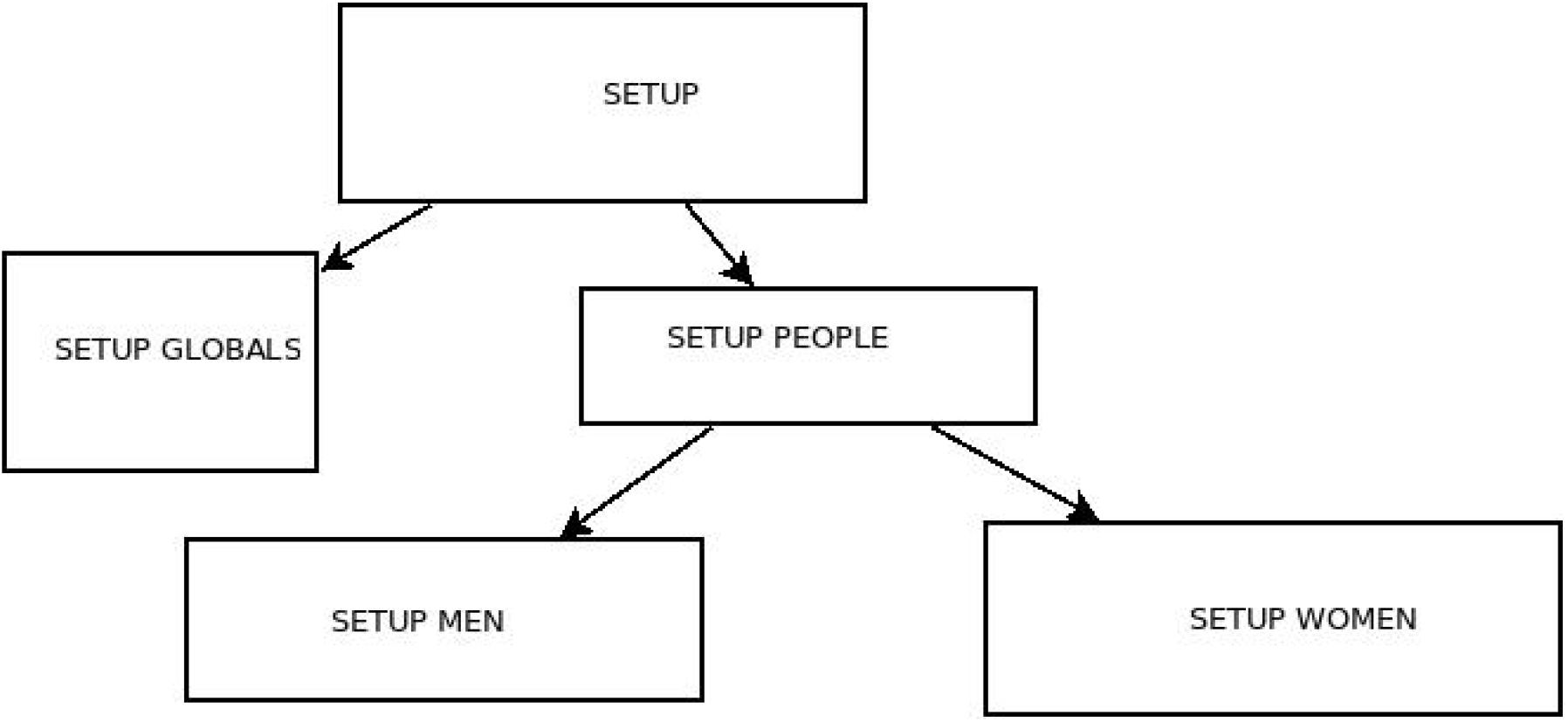
Setup method diagram.This initializes the simulation variables agent and global ones. Each variable initializes on random values near global ones.

The **Go** method dictate in which order the agent take action:

1. Move
2. if it has no couple, look for one.
3. if some threshold reaches, leave couple.
4. if it has a couple and is infected,
5. Screen-test with some probability.
6. If its infected and is a woman, and some threshold reaches with some probability *p* get sick and with probability 1 − *p* clearance

A diagram for this method is depicted in Figure 3

**Figure 3:**
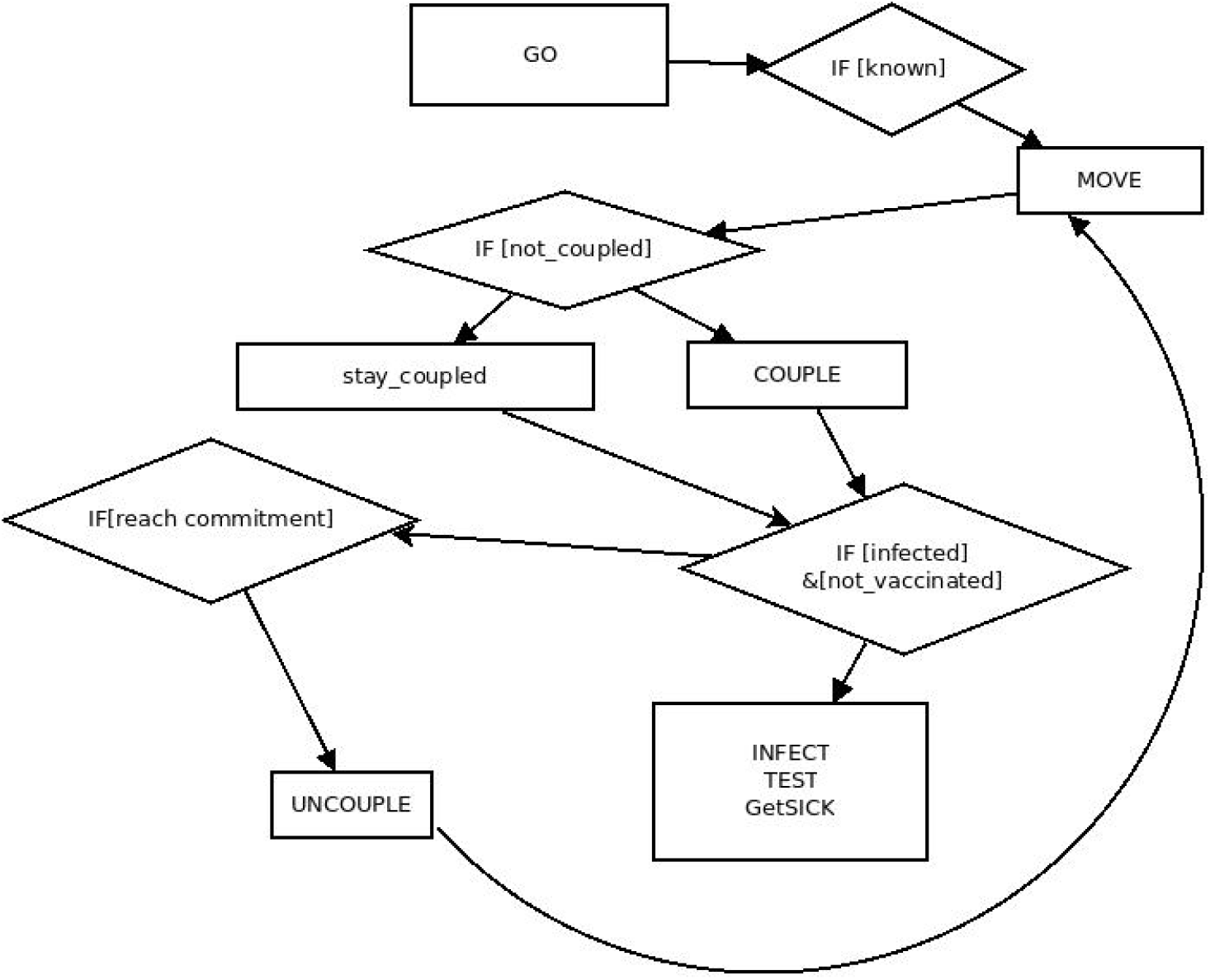
Go method diagram. This shows the actions taken for each agent at a single step of the simulation.

Two methods **toCouple** and **toInfect** define the key-steps on infection dynamics. In other models in the literature authors tend to simplify the couple “algorithm” making it full-random or full-effective or homogeneously distributed. Here we propose that there are individuals who are more effective to find a couple, that there is a biased process to find a couple. The coupling algorithm proposed is depicted on Figure 4

**Figure 4:**
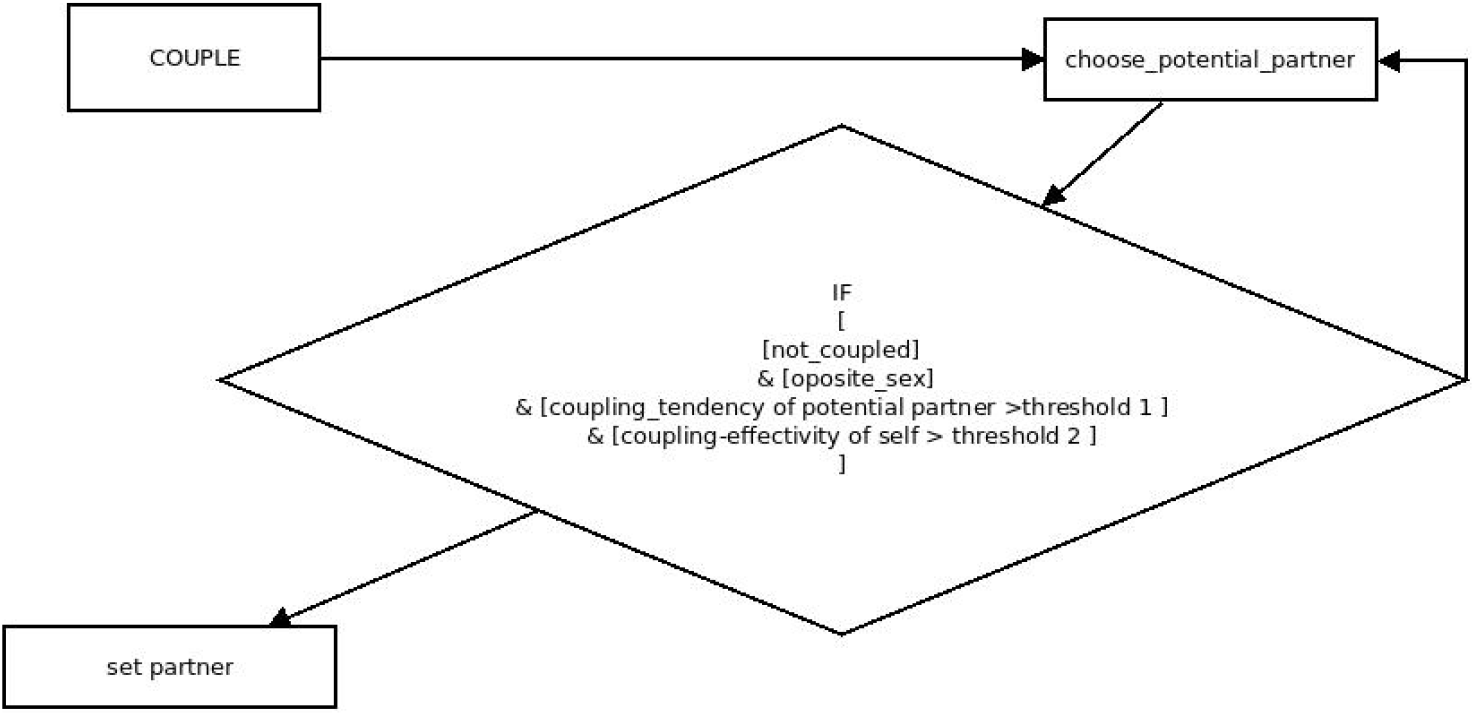
toCouple function diagram. This is the coupling algorithm for agents in HPV-VaC model, based on the principle that there exists a “natural” ability to find a new couple.

We set three different numerical experiments, for this work we have considered that there was no reason for explore different scenarios for the coupling tendency, although in [26] therEvidence and Impact of Human Papillomavirus Latencye is a comprehensive exploration of various agent-parameters. For this we have explored just three scenarios for average couple life-time for 14, 26 and 52 weeks. We have considered. And we have found on the trial that there was a greater coupling tendency for vaccinated women (almost twice). Coupling tendency is set on average 9 and promiscuity is normally distributed, but for vaccinated women are twice probably promiscuous according to data obtained in our population study (Table 2). We have not considered vaccinated males, for such an exploration refer to [26].

**Table 2:** Numerical experiments

| Experiment | Average couple life time (Weeks) | Average coupling tendency |
| --- | --- | --- |
| Experiment 1 | 14 | 9 |
| Experiment 2 | 26 | 9 |
| Experiment 3 | 52 | 9 |

## 3 Results

Our study population consisted of 18 HPV-vaccinated Mexican women aged 19-28 years (mean age, 23 years). We detected eight HPV-positive (44%) samples and ten HPV-negative (56%) samples. In two samples we detected a single infection with HPV16 type, in one sample we detected a multiple infection with HPV6, 61, 66 and 70 types, in one sample, we detected a multiple infection with HPV58 and 59 types, in one sample we detected a multiple infection with HPV56 and 66 types, in one sample we detected a multiple infection with HPV31, 53, and 81 types, in one sample we detected single infection with HPV71 type and in one sample we detected a single infection with HPV70 type. In summary, in four samples we detected a single infection and in four samples we detected multiple infections. On the other hand, in three samples we detected vaccine-HPV (16 or 6 types) and in five samples we detected non-vaccine HPV (others hr HPV types) (Figure 5 and Figure 6).

**Figure 5:**
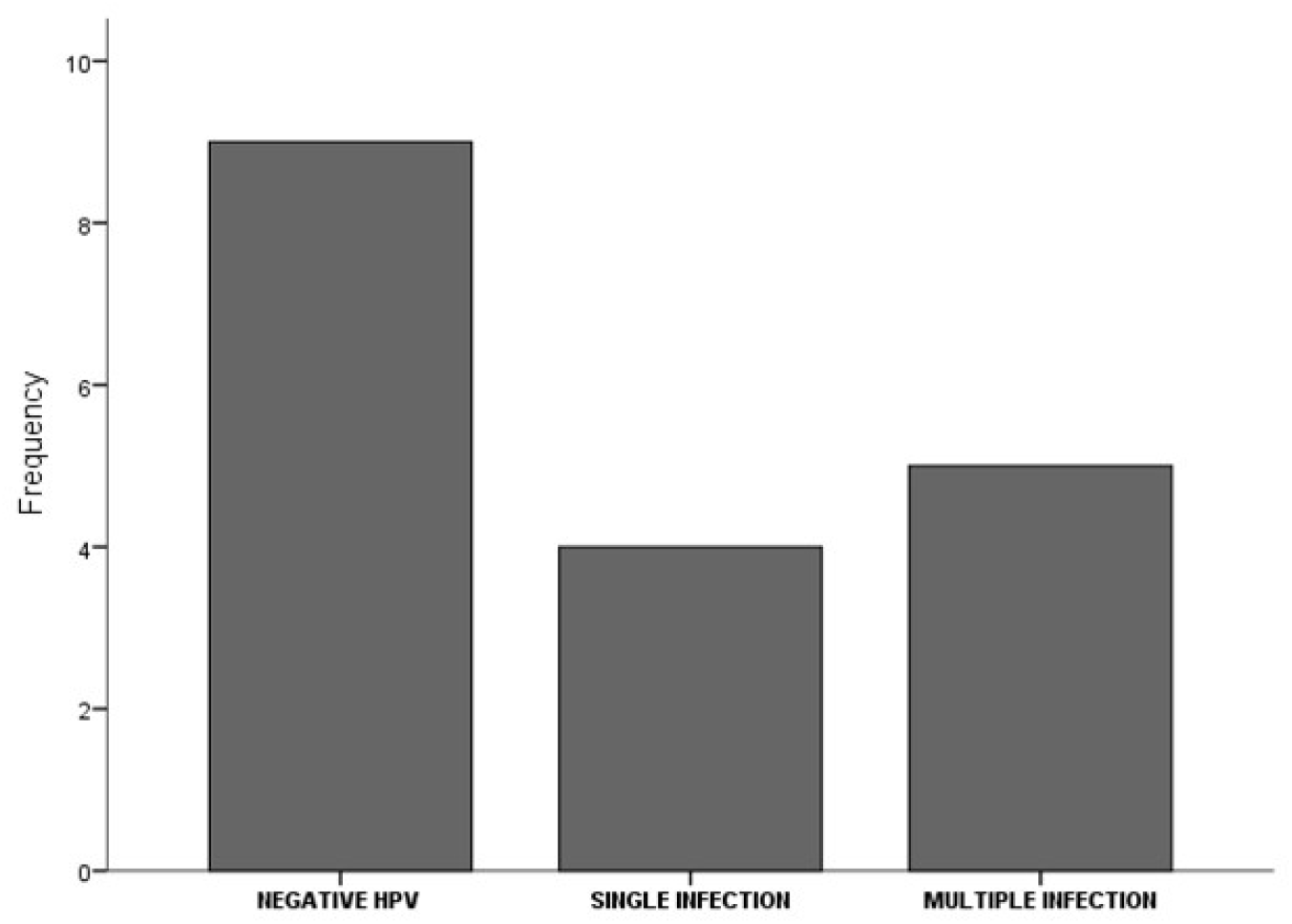
Frequency of simple, multiple and negative infections.

**Figure 6:**
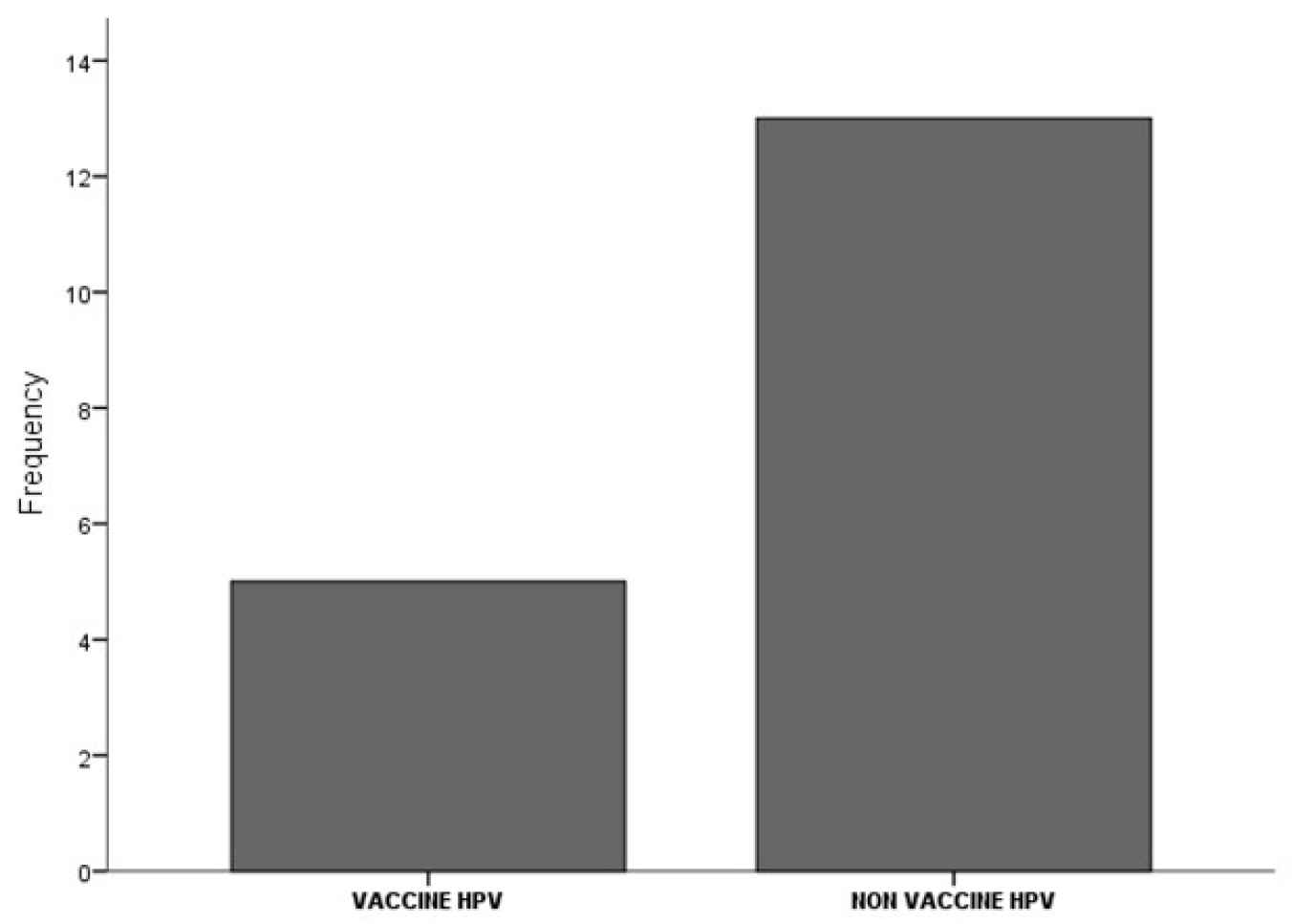
Frequency of vaccine HPV and non vaccine HPV detected in the analyzed samples.

The statistical association of the patient’s clinical variables with vaccine and non-vaccine HPV types were calculated. These data were obtained by questionnaire of interview and we included variables reported as important in HPV infections such as age, years of schooling, number of pregnancies, number of sexual partners, condom use, alcohol and smoking. Statistical analysis revealed no association between clinical variables and the HPV types detected in the samples (Tables 3 and 4). These results could suggest no association between vaccine and non-vaccine HPV types with some clinical variables.

**Table 3:** Association between HPV types and clinical variables

|  | Vaccine HPV | Non vaccine HPV | Bootstrap p value |
| --- | --- | --- | --- |
| Age |  |  |  |
| $\leq 25$ | 2 | 4 | 0.5380 |
| $> 25$ | 2 | 1 | |
| Years of schooling |  |  |  |
| $\leq 12$ | 1 | 1 | 0.7138 |
| $> 12$ | 3 | 4 | |
| Number of pregnancies |  |  |  |
| $< 1$ | 3 | 3 | 0.6660 |
| $\geq 1$ | 1 | 2 | |
| Number of sexual partners |  |  |  |
| $\leq 1$ | 0 | 1 | 0.7798 |
| $> 1$ | 4 | 4 | |
| Condom use |  |  |  |
| Yes | 1 | 2 | 0.6641 |
| No | 3 | 3 |  |
| Alcohol |  |  |  |
| Yes | 4 | 3 | 0.4140 |
| No | 0 | 2 |  |
| Smoking |  |  |  |
| Yes | 3 | 1 | 0.2903 |
| No | 1 | 4 |  |

**Table 4:** Association between HPV status and clinical variables

|  | HPV Positive | HPV Negative | Bootstrap p value |
| --- | --- | --- | --- |
| Age |  |  |  |
| $\leq 25$ | 6 | 5 | 0.6189 |
| $> 25$ | 3 | 4 | |
| Years of schooling |  |  |  |
| $\leq 12$ | 2 | 3 | 0.6237 |
| $> 12$ | 7 | 6 | |
| Number of pregnancies |  |  |  |
| $< 1$ | 6 | 8 | 0.4847 |
| $\geq 1$ | 3 | 1 | |
| Number of sexual partners |  |  |  |
| $\leq 1$ | 1 | 1 | 0.7120 |
| $> 1$ | 8 | 8 | |
| Condom use |  |  |  |
| Yes | 3 | 3 | 0.6503 |
| No | 6 | 6 |  |
| Alcohol |  |  |  |
| Yes | 7 | 8 | 0.6383 |
| No | 2 | 1 |  |
| Smoking |  |  |  |
| Yes | 4 | 3 | 0.6203 |
| No | 5 | 6 |  |

On the other hand, the results of numerical experiments of the agent-based model described above shows that there is a strong possibility that some strain of the virus will emerge stronger in a period of 10 years after the application of 5-valent vaccine on the first cohort of women (Figure 8 and Figure 9). The network of sexual contacts of agents in the model shows an increasing number of non-vaccine type HPV cases after vaccination, suggesting that high vaccination rate has driven a strain replacement after the first cohort of vaccinated women. In Figure 7 it’s depicted the plot for infections by distinct virus types and vaccinated women.

**Figure 7:**
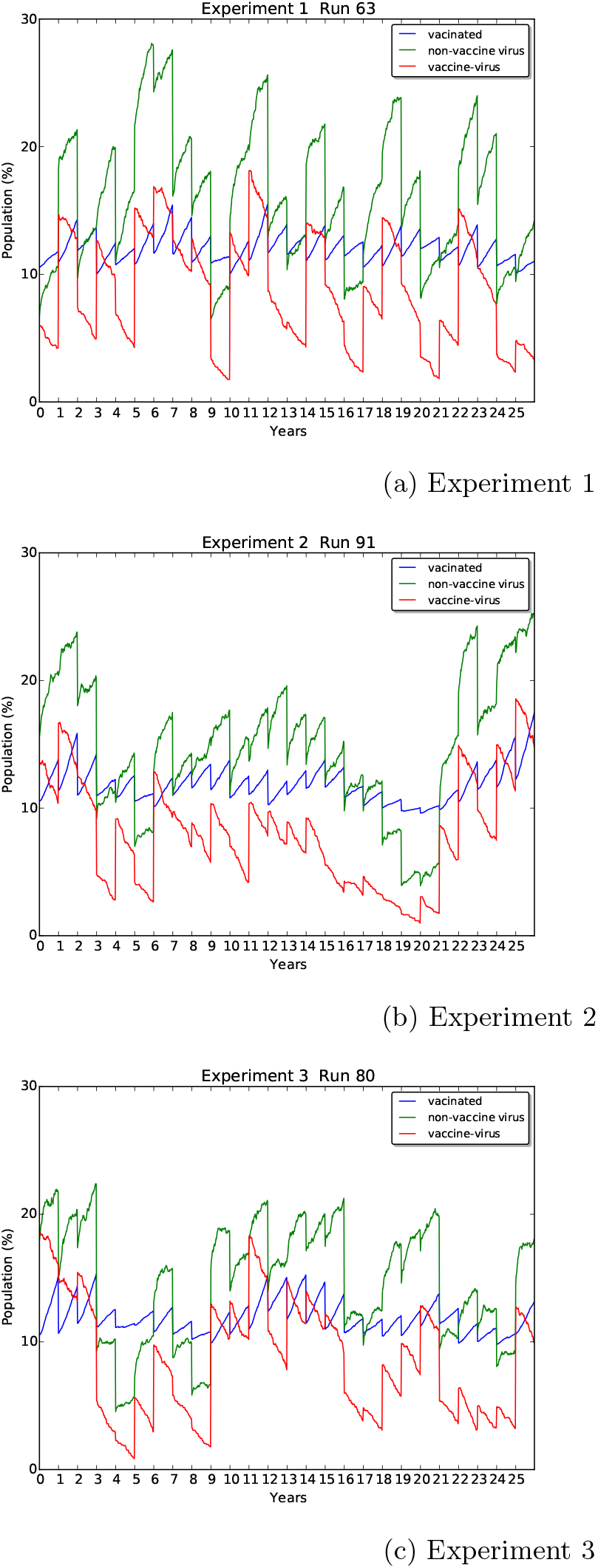
Three scenarios for Agent-Based model. The blue line represents vaccinated population, the red line represents infections by vaccine-type viruses, and the green line represents infections by non-vaccine-type viruses. In three cases one can see at least a spike afte4 approximately 10 years from vaccine application.

**Figure 8:**
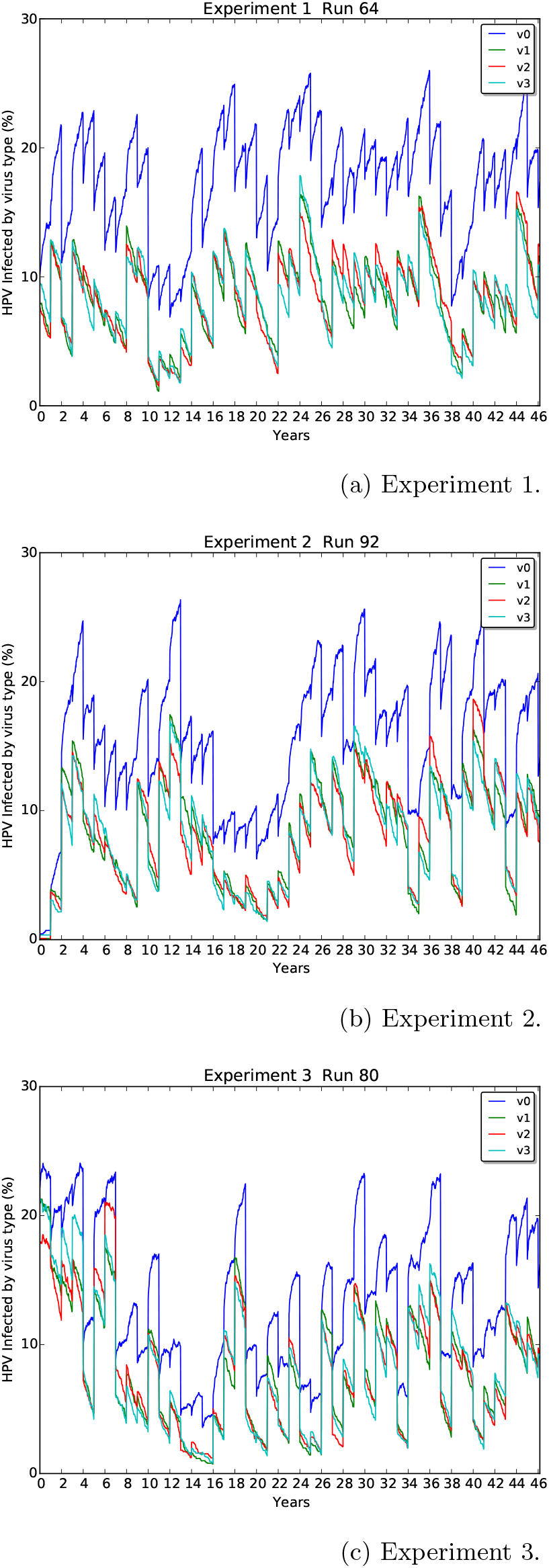
Time Series for infected population by four different families of HPV on three numerical experiments

**Figure 9:**
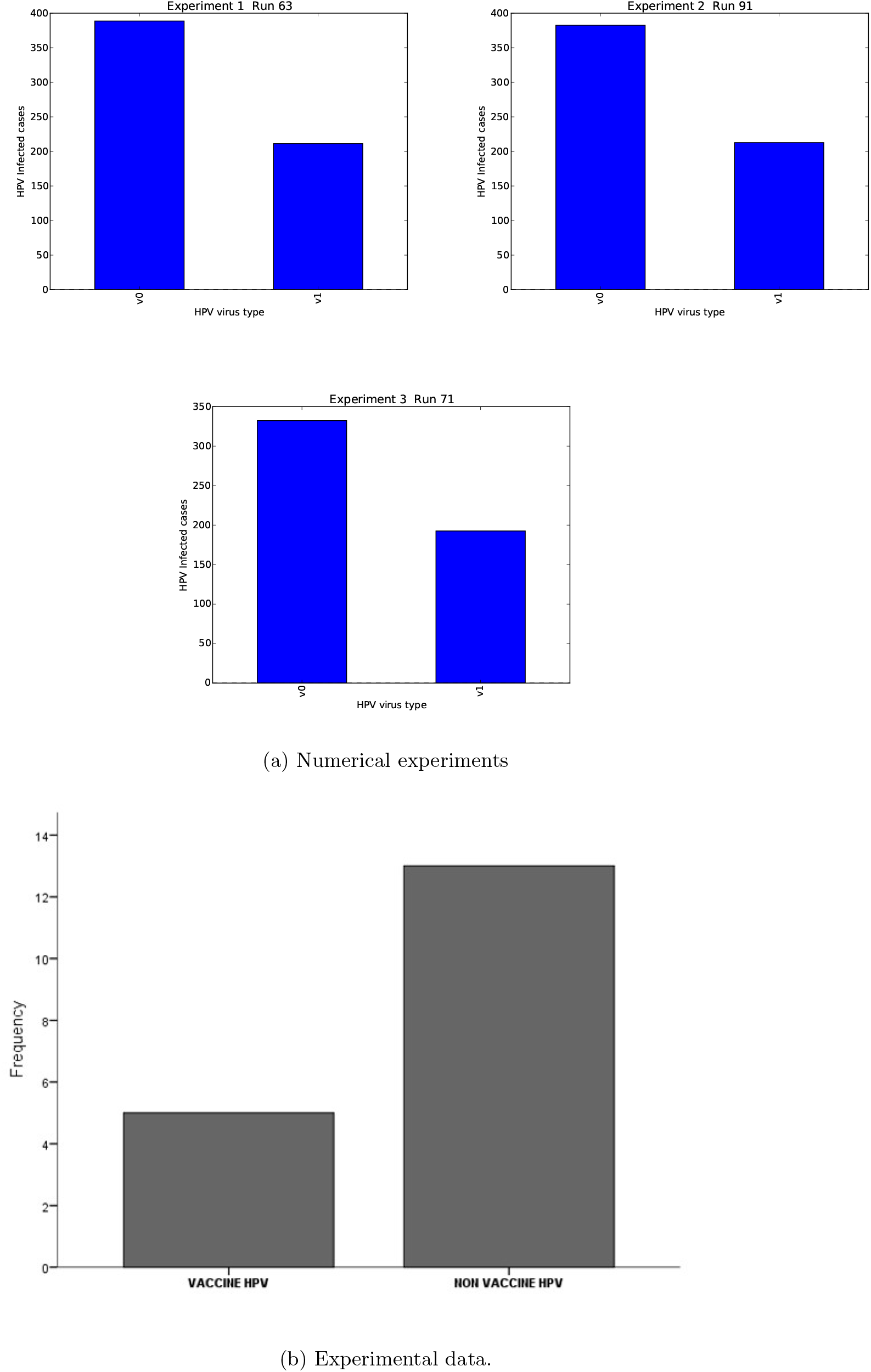
Bar plot for frequency of vaccine and non-vaccine type infections. Numerical experiments and experimental data obtained in pap test

## 4 Discussion

Vaccines are one of the most effective prophylactic measures to prevent infectious diseases. Thus, vaccines have increased human life expectancy and quality of life. In this respect, the introduction of the first vaccine against HPV in 2006 has allowed the decrease of infection of some HPV types. By October 2014, 68 countries had already implemented a vaccination program to prevent HPV infection [15]. HPV types 16 and 18 account for approximately 70% of cases of invasive CC, as well as a high proportion of anogenital cancer and a lower percentage of head and neck cancer, this fact led to the development of the vaccine against these two genotypes [16]. Recently, in some countries, two vaccines were introduced for the prevention of HPV16 and 18 infections: Cervarix®, a bivalent vaccine for HPV16 and 18, and Gardasil®, a tetravalent vaccine for HPV 16, 18, 6, and 11, the latter two HPV types,although have not high oncogenic risk, were included for prevent genital warts. Both vaccines provide prevention against HPV16 and 18 infections [15, 17]. In Mexico, the recommended strategy includes primary prevention with HPV vaccination for girls aged 9-14 years (before sexual initiation) [18].

Since application of the HPV vaccine, the incidence of HPV16/18 infections decreased by 64% in women younger than 20 years, while in women 20-24 years of age, the incidence of HPV16/18 decreased by 31%. Other genotypes that have decreased in incidence are HPV 31, 33 and 45 by 28%, indicating evidence of cross-protection in the population. With regard to genotypes not covered by the vaccine, these increased slightly [15]. These data indicate a possible association between the effectiveness of the vaccine and the age of the woman on the vaccine application. The vaccine is recommended in women prior to the initiation of sexual activity. The vaccine is not effective in women previously exposed to infection. When women are sexually active, they are more likely to have already been exposed to infection. The women participating in the present study were vaccinated before sexual initiation. However, at present they have started their sexual life and potentially exposed to the infection by vaccine and non-vaccine HPV. Some authors suggest that vaccinated patients might think that they have no risk of HPV infections [13].

Several epidemiological studies report the increase in non-vaccine infections in HPV-vaccinated women and a significant decrease in vaccine HPV16 and 18, and other HPV types related to vaccine HPV such as HPV31, 33, and 45 (suggesting cross-protection). On the other hand, a study reports a substantial fall in vaccine HPV types in HPV-vaccinated women and a lower prevalence of vaccine HPV types in unvaccinated women, suggested herd immunity and a possible indication of cross-protection against HPV types related to vaccine HPV types in HPV-vaccinated women [4]-[7]. In a recent study, Tota et al. have pointed that a type of replacement may occur, especially if different HPV types competitively interact during infection, but HPV type replacement does not occur among vaccinated individuals within four years and that is not likely to occur in vaccinated populations [19]. In this context, multiple infections have been frequently reported [20, 21]. In our study, 64% of the positive samples presented some multiple infection with at least two HPV types. In prior studies, this phenomenon was not easy to detect because the detection systems utilized had no way of determining it. By employing CLART® HPV2, we were able to demonstrate a high percentage of multiple infections (Figures 1,5 and 6).

On the other hand, the phylogenetic HPV classification shows that distributions of vaccine HPV are found in *α*9 and *α*7 species [22]. The HPV types of these species are genetically related, and epidemiological studies demonstrate a decrease by means of cross-protection. However, species α11, *α*5, and *α*6 are non-vaccine HPV and are considered hr oncogenic types. The increase in the incidence of HPV types of these species could support the hypothesis of VPSR in the case of the HPV vaccine. However further studies are required to support this hypothesis.

The use of SIR models in the study of the dynamics of HPV infection has helped to the understanding of the impact of HPV vaccination. Our previous model considers two genotypes: one that include the hr HPVs 16 and 18 or vaccine HPV and other including other hr HPV or non-vaccine HPV in our previous SIR model the non-vaccine HPV types increase after the application of the vaccine [9]. According to ecological notions, a population will expand its niche once another population is removed from a shared environment. In this context Murall et al. developed a mathematical model that represent multiple HPV infections and reported that a type replacement remains viable if nonvaccine HPV’s are not cross-reactive with the vaccine [23]-[25]. Based on the present results, the experimental data obtained in a group of HPV-vaccinated Mexican women were used for to normalize our previous model and confirm the dynamics of HPV infections after vaccination. This process of normalization includes the experiment

Agent-Based modeling constitutes a theoretical approach to understand the dynamics of complex systems such as epidemic systems. We have included the “HPV-VaC” model, calibrating the system parameters with the data obtained in the present study. In his Master’s thesis, Cabrera-Becerril has suggested [26] that current vaccination schemes are not totally effective. In that work, several possible scenarios for the interaction of the population were explored under three vaccination schemes. In this work we have reduced the scenarios to only three thanks to the use of experimental data to calibrate the model. As a result of the numerical experiments we have verified the existence of a strain replacement induced by the vaccine. In addition, there is a possible increase in cases of high-risk oncogenic HPV, approximately 10 years after the application of the vaccine to the first cohort of women.

## Data Availability

All data is publicly available

https://github.com/acb100cias/HPV

## 5 Acknowledgments

RP wishes to thank PRODEP-SEP 6986 for the support. The authors also wish to thank Hugo Castelán and Vladimir Rivera for his constructive comments and suggestions. This work was partially supported by UNAM-PAPIIT grant IN108318.

## 6 Competing interests

The authors have declared that no competing interests exist.

## 8 Author Contributions

Conceptualization: RP. Methodology in Silico: AC and CV. Methodology Experimental: RA, PR, KT and DM. Biological Samples: MC and SJ Supervision: PM. Validation: SD and MS. Writing – original draft: RP.

## Notes

### Competing Interest Statement

The authors have declared no competing interest.

### Funding Statement

Raul Peralta wishes to thank PRODEP-SEP 6986 for the support. This work was partially supported by UNAM-PAPIIT grant IN108318.

### Author Declarations

This study was approved by the Local Research Committee of University Health Center of the Morelos State University (UAEM).

## References

[1] Ma L, Cong X, Shi M, Wang XH, Liu HY, Bian ML. Distribution of human papillomavirus genotypes in cervical lesions. ExpTher Med. 2017; 13(2):535–541.

[2] Hernández-Ávila M, Torres-Ibarra L, Stanley M, Salmerón J, Cruz-Valdez A, Muñoz N, Herrero R, Villaseñor-Ruíz IF, Lazcano-Ponce E. Evaluation of the immunogenicity of the quadrivalent HPV vaccine using 2 versus 3 doses at month 21: An epidemiological surveillance mechanism for alternate vaccination schemes. Hum VaccinImmunother. 2016; 12(1):30–8.

[3] Durham DP, Poolman EM, Ibuka Y, Townsend JP, Galvani AP. Reevaluation of epidemiological data demonstrates that it is consistent with cross-immunity among human papillomavirus types. J Infect Dis. 2012; 206(8):1291–8.

[4] Cameron RL, Kavanagh K, Pan J, Love J, Cuschieri K, Robertson C, Ahmed S, Palmer T, Pollock KG. Human Papillomavirus Prevalence and Herd Immunity after Introduction of Vaccination Program, Scotland, 2009-2013. Emerg Infect Dis 2016;22(1):56–64.

[5] Mesher D, Soldan K, Howell-surname>Jones R, Panwar K, Manyenga P, Jit M, Gill ON. Reduction in HPV 16/18 prevalence in sexually active young women following the introduction of HPV immunisation in England. Vaccine. 2013;32(1):26–32.

[6] Tabrizi SN, Brotherton JM, Kaldor JM, Skinner SR, Liu B, Bateson D, McNamee K, Garefalakis M, Philips S, Cummins E, Malloy M, Garland SM. Assessment of herd immunity and cross-protection after a human papillomavirus vaccination programme in Australia: a repeat cross-sectional study. Lancet Infect Dis. 2014;14(10):958–966.

[7] Söderlund-Strand A, Uhnoo I, Dillner J. Change in population prevalences of human papillomavirus after initiation of vaccination: the high-throughput HPV monitoring study. Cancer Epidemiol Biomarkers Prev. 2014; 23(12):2757–64.

[8] Covert C, Ding L, Brown D, Franco EL, Bernstein DI, Kahn JA. Evidence for cross-protection but not type-replacement over the 11 years after human papillomavirus vaccine introduction. Hum Vaccin Immunother. 2019;15(7–8):1962-1969. doi: 10.1080/21645515.2018.1564438. Epub 2019 Feb 20. PubMed PMID: 30633598.

[9] Peralta R, Vargas-De-León C, Cabrera A, Miramontes P. Dynamics of high-risk nonvaccine human papillomavirus types after actual vaccination scheme. Comput Math Methods Med. 2014; 2014: 542923.

[10] Hodgkin AL, Huxley AF (August 1952). “A quantitative description of membrane current and its application to conduction and excitation in nerve”. The Journal of Physiology. 117 (4): 500–44.

[11] J Clin Epidemiol. 1996 Nov;49(11):1225–31. Advantages and disadvantages of using artificial neural networks versus logistic regression for predicting medical outcomes. Tu JV

[12] Gravitt PE. Evidence and impact of human papillomavirus latency. Open Virol J. 2012;6:198–203. doi: 10.2174/1874357901206010198. Epub 2012 Dec 28. PubMed PMID: 23341855; PubMed Central PMCID: PMC3547385.

[13] Ding L, Widdice LE, Kahn JA. Differences between vaccinated and unvaccinated women explain increase in non-vaccine-type human papillomavirus in unvaccinated women after vaccine introduction. Vaccine. 2017 Dec 19;35(52):7217–7221. doi: 10.1016/j.vaccine.2017.11.005. Epub 2017 Nov 21. PubMed PMID: 29169890; PubMed Central PMCID: PMC5712261.

[14] Naud PS, Roteli-Martins CM, De Carvalho NS, Teixeira JC, de Borba PC, Sanchez N, Zahaf T, Catteau G, Geeraerts B, Descamps D. Sustained efficacy, immunogenicity, and safety of the HPV-16/18 AS04-adjuvanted vaccine: final analysis of a long-term follow-up study up to 9.4 years post-vaccination. Hum Vaccin Immunother. 2014;10(8):2147–62. doi: 10.4161/hv.29532. PubMed PMID: 25424918; PubMed Central PMCID: PMC4896780.

[15] Harper DM, DeMars LR. HPV vaccines - A review of the first decade. Gynecol Oncol. 2017; 146(1):196–204.

[16] Jiang S, Dong Y. Human papillomavirus and oral squamous cell carcinoma: A review of HPV-positive oral squamous cell carcinoma and possible strategies for future. Curr Probl Cancer. 2017; doi:10.1016/j.currproblcancer.2017.02.006

[17] Castle PE, Maza M. Prophylactic HPV vaccination: past, present, and future. Epidemiol Infect. 2016; 144(3):449–68

[18] Lazcano-Ponce E, Allen-Leigh B. Innovation in cervical cancer prevention and control in Mexico. Arch Med Res. 2009; 40(6):486–92.

[19] Tota JE, Struyf F, Merikukka M, Gonzalez P, Kreimer AR, Bi D, Castell-sague X, de Carvalho NS, Garland SM, Harper DM, Karkada N, Peters K, Pope WA, Porras C, Quint W, Rodriguez AC, Schiffman M, Schussler J, Skinner SR, Teixeira JC, Hildesheim A, Lehtinen M; Costa Rica Vaccine Trial and the PATRICIA study groups.Evaluation of Type Replacement Following HPV16/18 Vaccination: Pooled Analysis of Two Randomized Trials. J Natl Cancer Inst. 2017 Jan 28;109(7). pii: djw300. doi: 10.1093/jnci/djw300. Print 2017 Jan.

[20] Choi YJ, Park JS. Clinical significance of human papillomavirus genotyping. J Gynecol Oncol. 2016; 27(2):e21.

[21] Romero-Morelos P, Uribe-Jiménez A, Bandala C, Poot-Vélez A, Ornelas-Corral N, Rodríguez-Esquivel M, Valdespino-Zavala M, Taniguchi K, Marrero-Rodríguez D, López-Romero R, Salcedo M. Genotyping of the human papilloma virus in a group of Mexican women treated in a highly specialist hospital: Multiple infections and their potential transcendence in the current vaccination programme. Med Clin (Barc). 2017; doi:10.1016/j.medcli.2017.02.021

[22] Burk RD, Chen Z, Van Doorslaer K. Human papillomaviruses: genetic basis of carcinogenicity. Public Health Genomics. 2009; 12(5–6):281-90.

[23] Elbasha EH, Dasbach EJ, Insinga RP. A multi-type HPV transmission model. Bull Math Biol. 2008;70(8):2126–76.

[24] Insinga RP, Dasbach EJ, Elbasha EH. Epidemiologic natural history and clinical management of Human Papillomavirus (HPV) Disease: a critical and systematic review of the literature in the development of an HPV dynamic transmission model. BMC Infect Dis. 2009;9:119.

[25] Murall CL, McCann KS, Bauch CT. Revising ecological assumptions about Human papillomavirus interactions and type replacement. J Theor Biol. 2014;350:98–109.

[26] Cabrera-Becerril, Augusto. Eficacia de la vacuna contra VPH: Un modelo basado en agentes. Masters degree Thesis http://132.248.9.195/ptd2017/junio/0760137/Index.html

[27] Cabrera-Becerril, Augusto. **ABM_VaC_min v0.3**. https://github.com/acb100cias/HPV

[28] Wilensky, U. (1999). NetLogo. http://ccl.northwestern.edu/netlogo/. Center for Connected Learning and Computer-Based Modeling, Northwestern University, Evanston, IL.

[29] Poleszczuk J., Macklin P., Enderling H. (2016) Agent-Based Modeling of Cancer Stem Cell Driven Solid Tumor Growth. In: Turksen K. (eds) Stem Cell Heterogeneity. Methods in Molecular Biology, vol 1516. Humana Press, New York, NY.

[30] Zhihui Wang, Joseph D. Butner, Romica Kerketta, Vittorio Cristini, Thomas S. Deisboeck, Simulating cancer growth with multiscale agent-based modeling, Seminars in Cancer Biology, Volume 30,2015, Pages 70–78.

